# Energetically costly functional network dynamics in cognitively impaired multiple sclerosis patients

**DOI:** 10.1101/2024.04.21.24306140

**Authors:** T.A.A. Broeders, M. van Dam, G. Pontillo, V. Rauh, L. Douw, Y.D. van der Werf, J. Killestein, F. Barkhof, C.H. Vinkers, M.M. Schoonheim

**Affiliations:** MS Center Amsterdam, Anatomy & Neurosciences, Vrije Universiteit Amsterdam, Amsterdam Neuroscience, Amsterdam UMC location VUmc, Amsterdam, The Netherlands; Radiology and Nuclear Medicine, Vrije Universiteit Amsterdam, Amsterdam Neuroscience, Amsterdam UMC location VUmc, Amsterdam, The Netherlands; Neurology,Vrije Universiteit Amsterdam, Amsterdam Neuroscience, Amsterdam UMC location VUmc, Amsterdam, The Netherlands; Psychiatry,Vrije Universiteit Amsterdam, Amsterdam Neuroscience, Amsterdam UMC location VUmc, Amsterdam, The Netherlands; Queen Square Institute of Neurology and Centre for Medical Image Computing, University College London, UK; Departments of Advanced Biomedical Sciences and Electrical Engineering and Information Technology, University of Naples “Federico II”, Naples, Italy; Amsterdam Public Health, Mental Health program, Amsterdam, The Netherlands; GGZ inGeest Mental Health Care, Amsterdam, The Netherlands

**Author notes:** **Corresponding author:** Tommy A.A. Broeders Department of Anatomy & Neurosciences, Amsterdam UMC, location VUmc, PO Box 7057, 1007 MB, Amsterdam, The Netherlands *Email:.

**Keywords:** multiple sclerosis, cognition, dynamic, network, states, control energy

## Abstract

**Background and objectives:** Patients with multiple sclerosis (MS) often experience cognitive impairment, and this is related to structural disconnection and subsequent functional reorganization. It is not clear how specific patterns of functional reorganization might make it harder for cognitively impaired (CI) MS patients to dynamically adapt how brain regions communicate, which is crucial for normal cognition. To identify patterns relevant to cognitive impairment in MS, we performed a temporally detailed analysis of connectivity state transitions and quantified the effort associated with such dynamic alterations.

**Methods:** Resting-state functional and diffusion MRI was acquired from 95 controls and 330 MS patients (mean disease duration: 15 years) in a cross-sectional design, of whom 86 were classified as CI (≥2/7 domains Z < −2) and 65 as mildly CI (≥2/7 domains Z < −1.5) based on the performance on an expanded Brief Repeatable Battery of Neuropsychological Tests. Four functional connectivity states were determined using *K*-means clustering of moment-to-moment co-fluctuations (i.e., edge time series), and the resulting state sequence was used to characterize the frequency of transitions between network configurations. The control energy associated with the transitions between states was then calculated using the structural network of each subject with network control theory.

**Results:** CI patients transition less frequently between connectivity states. Relative to the time spent in a particular state, CI patients specifically transition less from a weakly connected and highly modular state (i.e., the *weakly connected state*) to a more integrated state that featured strong involvement of the visual network (i.e., the *visual network state*), but more in the opposite direction. CI patients also required more control energy to transition between states.

**Discussion:** This study showed that it became more effortful for MS patients with cognitive impairment to dynamically change the organization of the functional network, providing an intuitive understanding of why these transitions occur less frequently in some patients. In particular, transitions between the *weakly connected state* and the more integrated *visual network state* were relevant for cognition in these patients. The findings highlight a possible underpinning of disturbed cognition in MS patients and also provide novel avenues for studying and possibly improving network dynamics.

## Introduction

Cognitive impairment is a highly debilitating symptom of multiple sclerosis (MS) that occurs in up to 65% of patients.^1^ In MS, neurodegeneration and neuroinflammation damage the central nervous system, giving rise to specific patterns of focal lesions and brain atrophy.^2^ These patterns can be detected using MRI, which is essential for diagnosis.^3^ However, the extent of MS-related structural brain damage observed using MRI is often not fully in line with clinical outcomes, particularly for cognitive impairment.^4^ In MS patients, damage to the white matter often disrupts the anatomical pathways between brain regions (i.e., structural connectivity) and this can also affect the communication between brain regions (i.e., functional connectivity).^5^ However, it is not fully understood how this may lead to cognitive impairment in MS.^6^

To that end, novel conceptual and mathematical tools were needed to describe how the MS brain is (dis)organized and why this leads to complex symptoms. In this push, network neuroscience has been crucial. In this field, the brain is represented as a graph consisting of brain regions (i.e., nodes) that are structurally and functionally connected (i.e., edges).^7^ Using this framework, it was learned that functional reorganization likely plays an important role in compensating for structural damage in the early stages of MS.^6^ Later in the disease, as structural damage accumulates, a critical threshold is passed after which the network cannot function properly.^8^ A key part of this loss of function seems to be the overload of a few highly-connected brain regions (i.e., hubs), such as regions in the default-mode network (DMN).^9^

A hub overload could leave the brain network as a whole less dynamically adaptable to cognitive challenges.^6^ However, these dynamic characteristics have often not been explicitly analyzed as most earlier studies averaged connectivity over time (i.e., static connectivity).^10^ This means that time-dependent patterns (i.e., dynamic functional connectivity) were neglected. Dynamic network alterations are integral brain processes that are relevant to cognitive functioning by themselves, for instance allowing the brain to transition between modes of segregated and integrated processing.^11^ A few more recent studies have observed disturbed network dynamics in MS patients with cognitive impairment, even without subjecting them to an explicit task (i.e., resting-state). For example, regions in the DMN, frontoparietal network (FPN) and visual network showed less connectivity dynamics.^12^ This has been interpreted as an indication that hubs can be stuck in an “overloaded” state.^6^ Brain network dynamics of cognitively impaired (CI) MS patients might be affected in non-hub regions as well,^13^ so it is important to look at dynamics of the functional network as a whole. Accordingly, recent studies applied a holistic model in which recurrent whole-brain connectivity patterns (i.e., “connectivity states”) were identified over time,^14^ showing that CI MS patients transitioned less fluidly between such states compared to cognitively preserved (CP) patients.^15^ Thus, less adaptability of the network might be particularly important for cognitive impairment in MS. Questions remain, including: which specific adaptations are important for cognitive impairment? Also, can structural network disturbances impede network adaptability?

Sensitivity to changes occurring on small temporal scales is needed to accurately characterize transitions between specific connectivity states. Nevertheless, connectivity dynamics have usually been captured using a sliding-window approach, which induces temporal blurring by computing correlations over windows of around a minute long.^16^ This issue can be ameliorated by temporally unwrapping correlation values and focusing on the resulting “edge time series”, which represent moment-to-moment co-fluctuations of regional brain activity. This approach makes it possible to disentangle brief events of high-amplitude co-fluctuations^16,17^ from non-events in between. Disentangling these could be useful, as both have been observed to relate to cognitive performance and may each provide unique insights.^18^

The interaction between brain structure and function previously related to cognitive impairment in MS,^19^ but an intuitive link to explain how the structural network could shape functional network dynamics has been missing. Network control theory can provide this link, by quantifying the control energy that is required to change the network configuration given the underlying structural network.^20^ Control energy has been likened to cognitive demand or mental load, so these measures quantify how effortful transitions between connectivity states are. Recent work showed that physically disabled MS patients required more control energy to transition between activity states,^21^ but it is unclear whether this can explain the disrupted functional network dynamics in patients with cognitive impairment.

Therefore, this work aimed to increase our understanding of the functional underpinnings of cognitive impairment in MS. This is done by characterizing framewise state transitions and computing the control energy required for these connectivity state transitions. We investigated 330 patients with MS and 95 healthy controls (HCs) and hypothesized that MS patients with cognitive impairment would remain “stuck” in (i.e., transition away less from) states featuring relatively high DMN connectivity. This pattern was expected to be explained by the energetic costs of making the transitions.

## Methods

### Participants

Cross-sectional imaging from the Amsterdam MS cohort was analyzed, including a total of 330 MS patients and 95 HCs with available functional and diffusion MRI data who were recruited between 2008 and 2012. Functional network dynamics has been described previously for these participants,^12^ but not yet in combination with diffusion MRI data. All patients were diagnosed with clinically definite MS according to the 2010 revised McDonald criteria.^3^ These patients were relapse-free and without steroid treatment for two months before participation, and have no history of a psychiatric and/or neurological disease besides MS. Age, sex and the highest obtained level of education were acquired from all participants and clinical data obtained from patients included: symptom duration, disease phenotype and treatment status. The Expanded Disability Status Scale (EDSS) was administered by a neurologist to characterize the level of physical disability. Fatigue was ascertained in a subset of patients (*N*=167) with the Checklist Individual strength-20 Revised. Approval for the study was acquired from the local institutional ethics review board. All participants provided written informed consent before participation.

### Neuropsychological assessment

An expanded Brief Repeatable Battery of Neuropsychological Tests was used on the same day as the MRI examination, as previously described.^22^ Performance on individual tests was grouped into seven cognitive domains and adjusted for age, sex and education effects in the HCs. Domains included executive functioning (concept shifting), verbal memory (selective reminding), information processing speed (symbol digit modalities), verbal fluency (word list generation), visuospatial memory (spatial recall), working memory (memory comparison) and attention (stroop colour-word). The scores of these domains were transformed to *z*-scores based on the distribution of HCs. These *z*-scores were averaged across domains to produce a summary value of average cognition, used to explore linear relationships (not classify cognitive groups). To classify cognitive groups, performance on all domains was compared to HCs, resulting in three groups in MS: CI, mildly CI (MCI), CP. Classification of CI patients was defined as scoring at least two standard deviations below HCs on two or more cognitive domains.^9^ Patients that were not defined as CI, but scored at least 1.5 standard deviations below HCs on two or more cognitive domains, were classified as MCI. All other patients were classified as CP.

### MRI acquisitions

All scans were acquired using a 3T MRI scanner (GE Signa-HDxt, Milwaukee, WI) with an 8-channel phased-array head coil. The protocol included a 3D T1-weighted fast spoiled gradient echo sequence (repetition time[TR]/echo time[TE]=7.8/3ms; inversion time=450ms; flip angle=12°; sagittal slice thickness=1.0mm; in-plane resolution=0.9×0.9mm), a 3D T2-weighted fluid-attenuated inversion recovery (FLAIR) sequence (TR/TE=8000/125ms; inversion time=2350ms; sagittal slice thickness=1.2mm; in-plane resolution=1.0×1.0mm), a resting-state fMRI echo planar imaging sequence (202 volumes; TR/TE=2200/35ms; flip angle=80°; axial slice thickness=3mm, contiguous; in-plane resolution=3.3×3.3mm) and a diffusion tensor imaging sequence using five volumes without directional weighting (b=0 s/mm^2^) and 30 with non-collinear diffusion gradients (b=1000s/mm^2^, TR/TE=13000/91ms, flip angle=90°, axial slice thickness=2.4mm, contiguous; in-plane resolution=2×2mm).

### Image preprocessing

#### Lesion detection and filling

White matter lesions of MS patients were automatically segmented on FLAIR images^23^ and the resulting lesion masks were linearly registered to T1-space to perform lesion filling.^24^

#### Functional preprocessing

The fMRI images of all 425 participants were preprocessed using the MELODIC pipeline (FSL 6, fmrib.ox.ac.uk/fsl), including the removal of the first two volumes, motion correction, brain extraction and 4mm Gaussian smoothing. Subsequently, ICA-AROMA (v0.4-beta)^25^ was used for automatic removal of residual motion artifacts. Then, regression of mean white matter and cerebrospinal fluid signal was performed, followed by high-pass temporal filtering, boundary-based registration to T1-space and co-registration and resampling to 4mm standard space.

#### Diffusion preprocessing

Complete diffusion MRI data was available for 420 participants. Preprocessing was performed using QSIPrep 0.14.3.^26^ This included denoising and correction for B1 field inhomogeneity, head motion and eddy currents. Then, a deformation field was estimated using a registration-based fieldmap-less approach and used to calculate an unwarped b0 reference.^27^ The unwarped diffusion data was then registered to the T1w volume with 2mm isotropic voxels. Fiber response functions and orientation distributions (FODs) were produced with an unsupervised multi-tissue method and subsequent intensity normalization.^28^

### Structural damage indicators

#### Volumetric measures

FreeSurfer 7.1.1 was used to automatically perform white-matter as well as cortical and deep gray matter segmentations on T1 weighted images.^29^ Lesion segmentation masks were used to determine white matter lesion volume.

#### White matter integrity

Fractional anisotropy (FA) was calculated for each voxel using DSI studio.^30^ FA maps were nonlinearly registered and projected onto an FA template skeleton, using the tract-based spatial statistics pipeline.^31^ The mean FA over the whole skeleton signified white matter integrity.

### Network generation

#### Functional networks

All 210 cortical regions from the Brainnetome atlas^32^ were combined with 14 deep gray matter (DGM) segmentations derived from FSL’s FIRST and transformed to fMRI-space. For visualization, all regions were assigned to one of seven cortical resting-state subnetworks^33^ based on maximum overlap. All DGM regions were combined into one distinct network. Only voxels that represented gray matter were included, whereas distorted resting-state fMRI signal was excluded from the atlas.^9^ Regions with less than 30% non-distorted voxels in more than 10% of participants were discarded (the bilateral orbitofrontal and nucleus accumbens). This resulted in 197 brain regions, from which nodal (i.e., regional) functional time series were extracted. We computed edge time series by transforming nodal time series to *z*-scores (using nodal means and standard deviations) and performing point-wise multiplication.^16,17^ Edge time series were used to determine a functional network for each frame in the scan (see Figure 1A).

**Figure 1.**
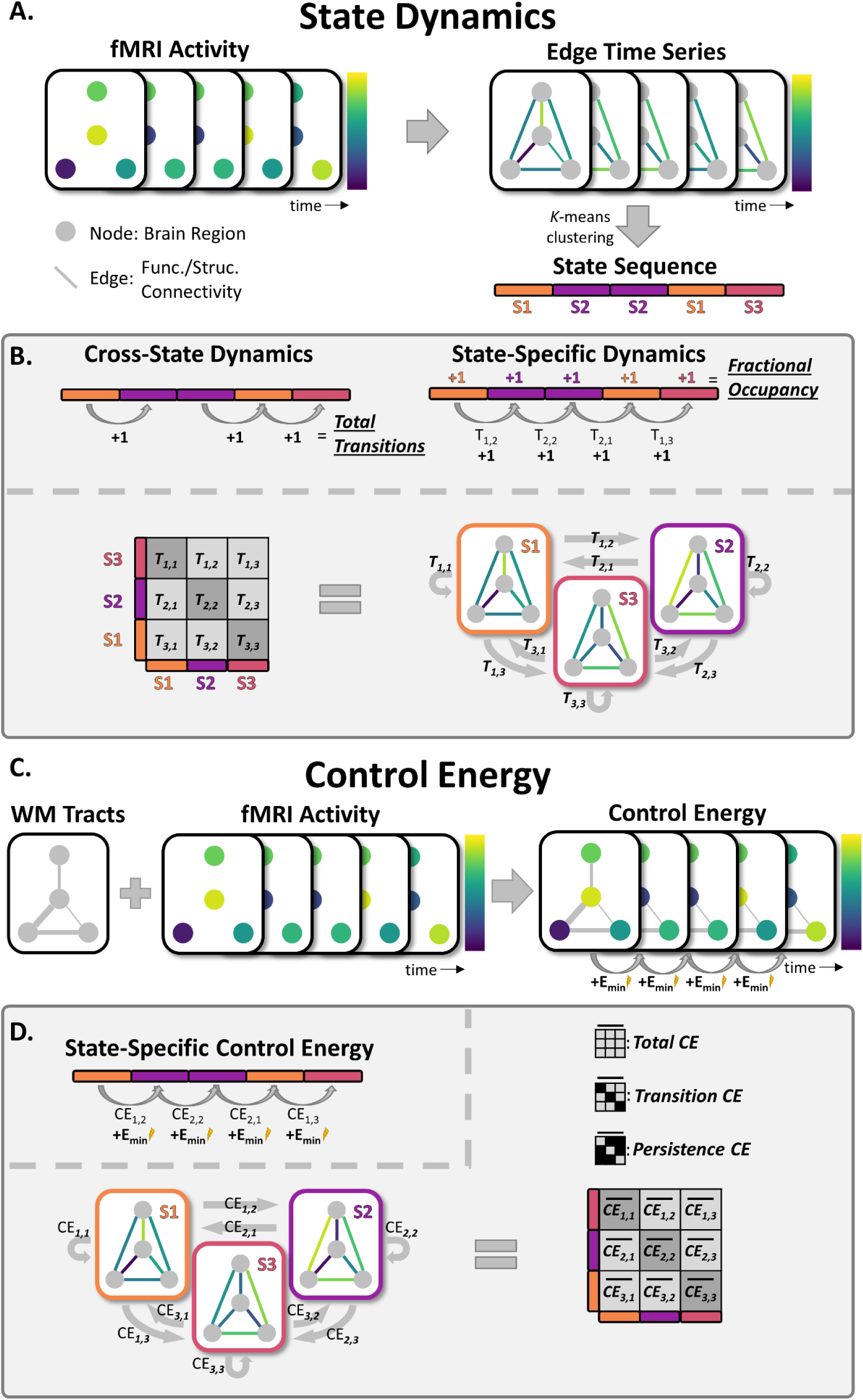
Illustration of the quantification of state dynamics and control energy. **A)** Functional MRI (fMRI) data was used to create edge time series, which reflected a functional network per time point. These networks were clustered using *K-*means clustering to define brain states (S1-S3 in this example; each represented by different colors). **B**) Cross-state dynamics were quantified using the total number of transitions, whereas fractional occupancy (i.e., fraction of time spent in each state) was used as state-specific measure and the transition probability (T_i,j:_ probability of transitioning from state *i* to state *j,* relative to the total transitions from *i*) as transition-specific measure. **C**) Information on the number of streamlines of white-matter (WM) tracts (based on diffusion MRI) was combined with fMRI data, to derive the minimum control energy (E_min_) that is required to transition between successive frames. **D**) The resulting E_min_ values were averaged over the same transitions (using the state sequence) to compute an energy transition matrix for each participant, so this matrix denotes the mean E_min_ required for each type of transition. The mean over the total matrix signified total control energy (CE), whereas the diagonal and off-diagonal reflected transition- and persistence CE.

#### Structural networks

Tractography was performed using MRTrix3 using the normalized white matter FODs, by applying iFOD2 probabilistic tracking to generate 10 million streamlines.^34^ Anatomical constraints were provided by a hybrid surface/volume segmentation.^35^ Finally, streamline weights were calculated using SIFT2^36^ and a 196×196 structural connectivity matrix was filled with the weighted number of streamlines connecting brain regions, using only regions in the functional network (excluding the cerebellum).

### State dynamics

#### State identification

Edge time series of all participants were concatenated and *k*-means clustering was performed to derive connectivity states.^14^ Clustering was performed in MATLAB for 2-7 clusters, with five replicates and using city block distance. The optimal number of states was derived using the elbow criterion, resulting in four states whose centroids (cluster median) represented robustly detected co-activation patterns. The resulting state sequence assigns each frame to a connectivity state. The organization of these states was characterized by computing global mean connectivity, global efficiency, modularity, and the eigenvector centrality per resting-state network of the centroids, using the brain connectivity toolbox.

#### State dynamics characterization

The *total number of transitions* across all states, as well as the *average fractional occupancy* (time spent in each state) and *relative transition probabilities* (probability of transitioning between and persisting within each individual state) were computed (see Figure 1B).^14^ In line with arguments made in previous research,^37^ the relative transition probabilities from states that were not visited were considered missing values in this work.

### Control energy

The nctpy Python toolbox was used to determine control energy based on network control theory (see eMethods 1 for details). The *minimum control energy* (*E_min_*) was calculated per brain region, reflecting the minimum external input that explains the observed change in brain activity (see Figure 1C). The sum across brain regions defined overall required energy for a transition. Averaging the *E_min_* from the same state transitions using information from the state sequence, resulted in a 4×4 control energy transition matrix per participant (see Figure 1D). Each transition in the transition matrix was transformed to a *z*-score based on the distribution of HCs for that transition. The average across the entire matrix determined the *total control energy* required across state transitions, representing overall energy inefficiency or energetic costs. The means of the diagonal values determined *persistence control energy*, denoting the costs of staying in the same state. The off-diagonal values were used for *transition control energy*, signifying the costs of transitioning to a different state. Some transitions were not observed in specific individuals, so the control energy for these transitions were regarded as missing values.

### Statistical analyses

SPSS 28 was used for all statistical analyses. All group comparisons (unless differently specified) were adjusted for age, sex and education. Education was based on the highest level attained and was binarized for analyses (higher vocational education or university yes/no). When the same analysis was performed across multiple states or transitions, multiple comparisons were taken into account using Bonferroni and corrected *p*-values were reported. An α-level of .05 was considered statistically significant. Normality of the dependent variables was inspected visually and using Kolmogorov-Smirnov tests.

Demographics and clinical variables were compared between the cognitive groups (HC, CP, MCI and CI) using chi-square (*X*^2^) tests of independence for categorical variables and ANOVAs for numeric variables (no adjustment for covariates). For group comparisons of all imaging measures, linear mixed models were used when the data was normally distributed and Quade’s non-parametric ANCOVA if not. Using this approach, differences in the total number of state transitions and the fractional occupancy of each specific state were compared between cognitive groups. Transition probabilities were investigated for states that showed differences between groups. Total control energy as well as transition and persistence control energy were compared between the cognitive groups. Then, the difference in control energy required to transition between specific states was evaluated. Finally, the connection between transition probability and control energy was investigated in relation to measures of structural damage and clinical indicators of MS using partial correlations.

## Results

### Demographics and clinical characteristics

Complete fMRI and neuropsychological assessments were available for 330 patients (mean age of 48 ± 11 years; 68% female) and 95 HCs (mean age of 46±10 years, 58% female). Across all patients, 179 (54%) were classified as CP (131 women, mean age of 46 ± 10 years), 65 (20%) as MCI (42 women, mean age of 49 ± 12 years) and 86 (26%) as CI (51 women, mean age of 51 ± 11 years). Cognitive groups differed on age, sex and educational level (see Table 1).

**Table 1.** Demographic, clinical and brain volumetric sample characteristics.

|  |  | Multiple Sclerosis (MS) |  |  |  |  |
| --- | --- | --- | --- | --- | --- | --- |
|  | HC (N=95) | CP (N=179) | MCI (N=65) | CI (N=86) | Test-statistic | *p*-value |
| **Demographics** |  |  |  |  |  |  |
| Male, *n* | 40 (42.1%) | 48 (26.8%) | 23 (35.4%) | 35 (40.7%) | $\chi^2=8.607$ | **0.035\*** |
| Age, *y* | 45.70 ± 10.35<sup>CI</sup> | 46.16 ± 10.35<sup>CI</sup> | 49.19 ± 12.15 | 51.21 ± 10.66<sup>HC,CP</sup> | $F=5.819$ | **<0.001\*** |
| Level of education<sup>‡</sup> | 6 (3)<sup>MCI,CI</sup> | 6 (2)<sup>MCI,CI</sup> | 4 (3)<sup>HC,CP</sup> | 4 (3)<sup>HC,CP</sup> | $F=7.035$ | **<0.001\*** |
| **Disease characteristics** |  |  |  |  |  |  |
| Symptom duration | - | 13.49 ± 7.83<sup>CI</sup> | 14.15 ± 8.15 | 17.17 ± 9.30<sup>CP</sup> | $F=5.799$ | **0.003\*** |
| Disease phenotype,<br>RRMS/SPMS/PPMS | - | 147<sup>CI</sup>/20<sup>CI</sup>/12<sup>MCI</sup> | 47/6<sup>CI</sup>/12<sup>CP</sup> | 49<sup>CP</sup>/25<sup>CP,MCI</sup>/12 | $\chi^2=26.106$ | **0.001\*** |
| Treatment,<br>Yes, *n* | - | 63 (35.2%) | 26 (40.0%) | 57 (33.7%) | $\chi^2=0.689$ | 0.709 |
| IFB/COP/NA/Other | - | 37/6/16/4 | 18/6/1/1 | 18/4/6/1 | $\chi^2=7.666$ | 0.264 |
| **Clinical Variables** |  |  |  |  |  |  |
| EDSS<sup>‡</sup> | - | 3 (2)<sup>CI</sup> | 3 (1.5)<sup>CI</sup> | 4 (2.75)<sup>CP,MCI</sup> | $F=25.360$ | **<0.001\*** |
| Cognitive function | 0.01 ± 0.48<sup>a</sup> | -0.19 ± 0.46<sup>a</sup> | -1.00 ± 0.34<sup>a</sup> | -1.90 ± 0.92<sup>a</sup> | $F=221.480$ | **<0.001\*** |
| **Brain Volume** |  |  |  |  |  |  |
| Cortical GM, *BPF* (%) | 32.28 ± 1.52<sup>CI</sup> | 31.99 ± 1.77<sup>CI</sup> | 31.69 ± 1.99<sup>CI</sup> | 30.59 ± 1.71<sup>a</sup> | $F=16.823$ | **<0.001\*** |
| Deep GM, *BPF* (%) | 3.72 ± 0.21<sup>a</sup> | 3.49 ± 0.28<sup>HC,CI</sup> | 3.39 ± 0.32<sup>HC,CI</sup> | 3.15 ± 0.35<sup>a</sup> | $F=60.572$ | **<0.001\*** |
| Lesion volume, *mL* | - | 10.28 (8.63)<sup>a</sup> | 14.05 (11.02)<sup>a</sup> | 22.09 (17.00)<sup>a</sup> | $F=21.126$ | **<0.001\*** |

*Note.* All values represent means and standard deviations for the continuous variables, but signify medians and the interquartile range (<sup>‡</sup>) or frequencies for categorical variables. Sample characteristics were compared between all groups. The level of education was based on the highest level of education attained. Brain lesion volume was transformed to milliliters (mL) for readability. Post-hoc pairwise comparisons were Bonferroni corrected (<sup>a</sup>=significantly different from all other groups, <sup>HC</sup>=significantly different from HC, <sup>CP</sup>=significantly different from CP, <sup>MCI</sup>=significantly different from MCI, <sup>CI</sup>=significantly different from CI). HC=healthy control, CP=cognitively preserved, MCI=mild cognitive impairment, CI=cognitive impairment, RRMS=relapsing-remitting MS, SPMS=secondary progressive MS, PPMS=primary progressive MS, IFB=interferon-beta, cop=copaxone, NA=natalizumab, EDSS=expanded disability status scale, GM=gray matter, BPF=brain parenchymal fraction, \**p*<0.05.

### State dynamics

#### State organization

Four recurring connectivity states were identified (see Figure 2A). *State 1* was moderately connected with high visual network and therefore described as the *visual network state* (see Figure 2B). *State 2* was strongly connected overall, thus termed the *highly connected state*. *State 3* showed moderate connectivity strength, in particular with a strongly connected VAN and therefore called the *VAN state*. *State 4* was weakly connected with high modularity and high DMN connectivity and described as the *weakly connected state*.

**Figure 2.**
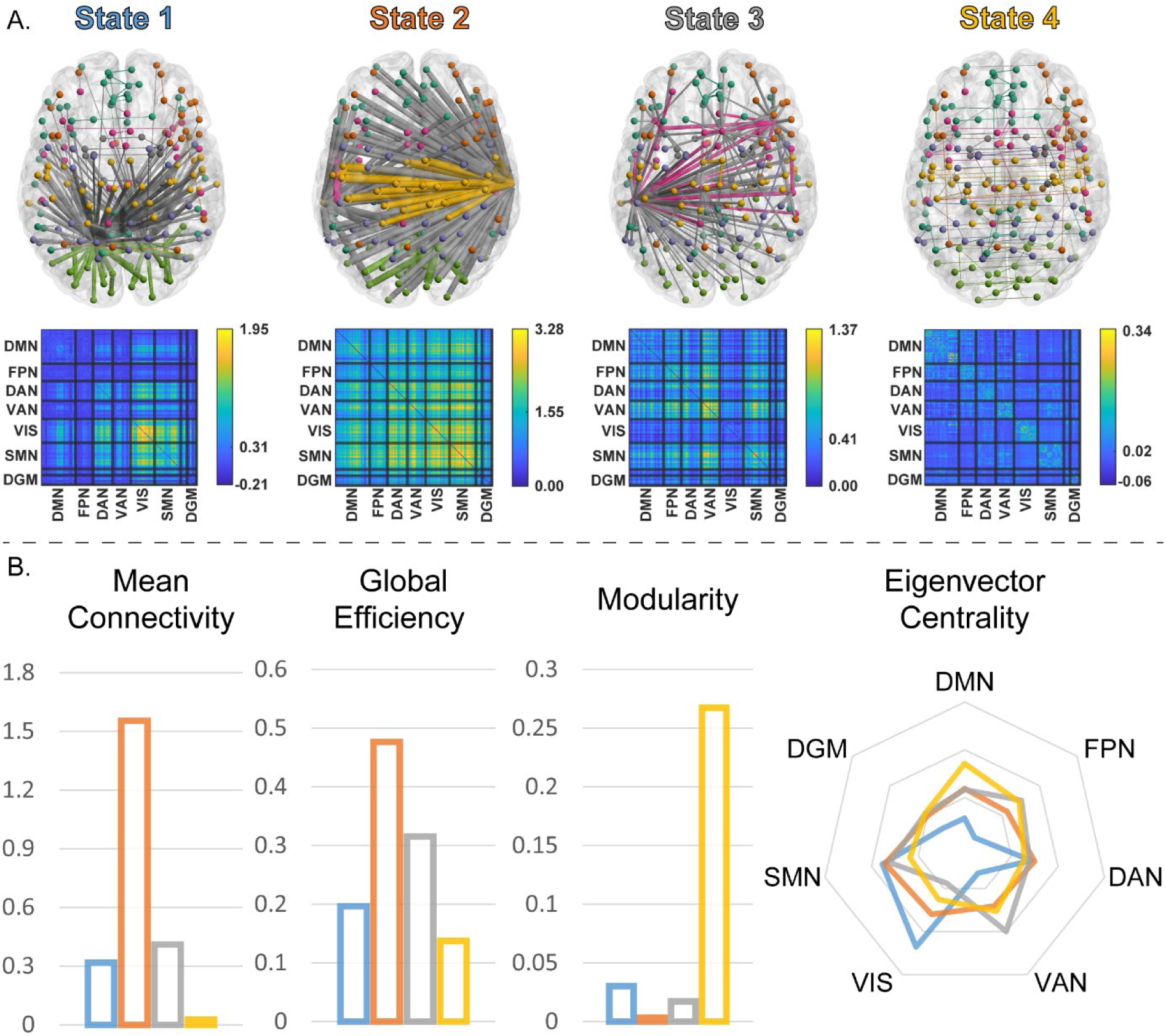
Brains state organization. Four states were identified using *K*-means clustering. **A**) The backbone of the network of the state (minimum spanning tree) is depicted, with the thickness indicating connection strength and the colored edges indicating within-network connections and gray edges between network connections. The corresponding connectivity matrices are depicted below. ***B***) Global connectivity strength, global efficiency, global modularity and the mean eigenvector centrality per resting-state network are portrayed per state centroid to help illustrate how states differed from each other. *Note. DMN=default-mode network, FPN=frontoparietal network, DAN=dorsal attention network, VAN=ventral attention network, VIS=visual network, SMN=sensorimotor network, DGM=deep gray matter*.

#### Total transitions

The total number of transitions was lower in CI compared to CP patients and HCs (see Figure 3 & Table 2 for statistical parameters). MCI patients also transitioned between states less frequently compared to HCs. Thus, dynamics became less fluid in impaired patients.

**Figure 3.**
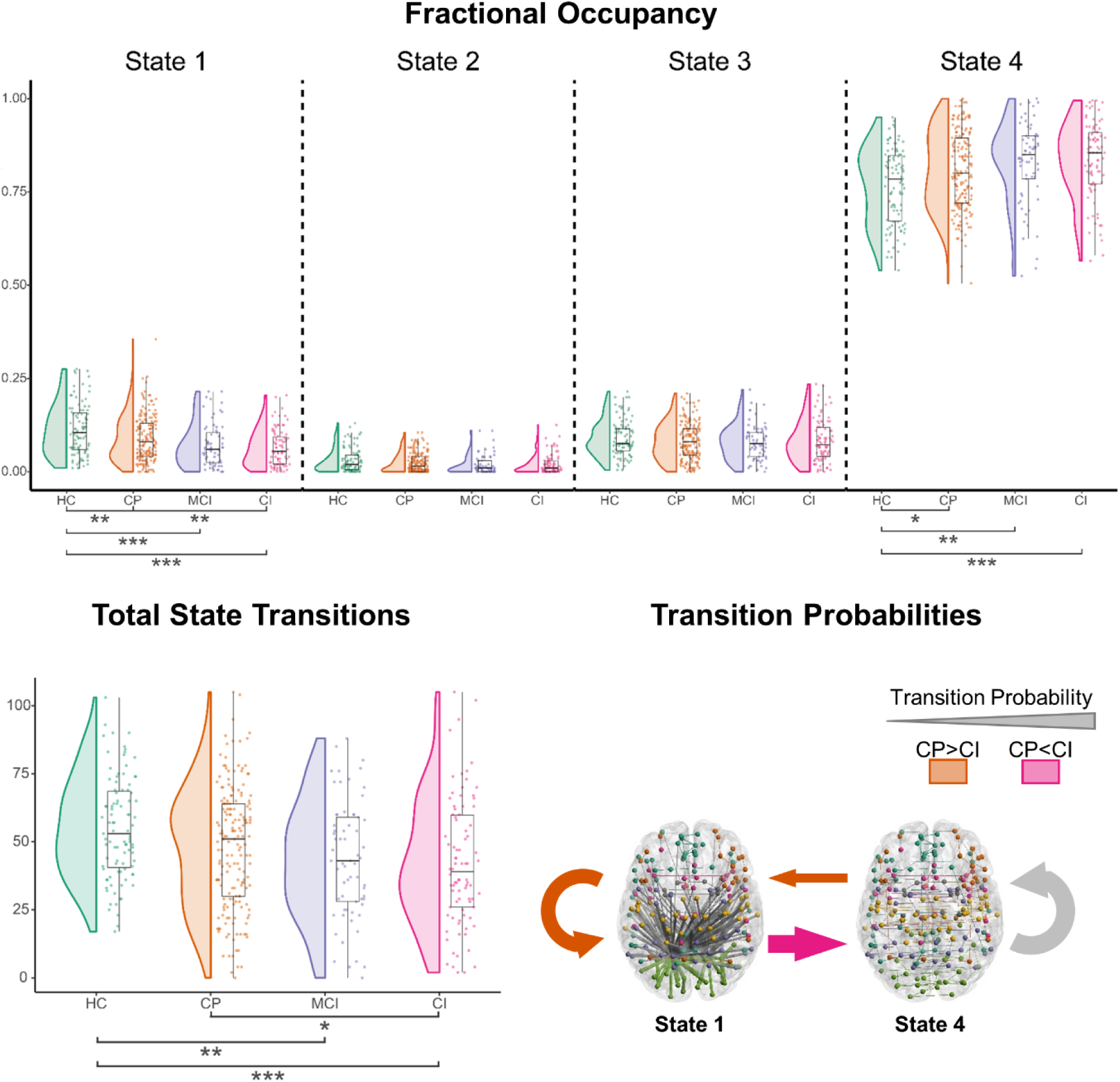
Brains state dynamics across cognitive groups. In CI patients, fewer transitions between brain states were observed compared to CP and HCs. These patients spend less time in *State 1* and more time in *State 4*. CI patients show a lower probability to move from *State 4* to *State 1* and stay there relative to preserved patients, whereas the transition from *State 1* to *State 4* was more likely. The thickness of the arrows on the bottom right indicates the relative probability of that transition occurring on average in HCs. *Note. HC=healthy control, CP=cognitively preserved, MCI=mildly impaired, CI=cognitively impaired, S1-S4=State 1- State 4, *p<0.05, **p<0.01, ***p<0.001*.

**Table 2.** State dynamics and control energy across cognitive groups.

| | Mean (±SD) | | | | Main: Group | | CI vs CP | |
| --- | --- | --- | --- | --- | --- | --- | --- | --- |
| | HC (N=59) | CP (N=134) | MCI (N=42) | CI (N=54) | F | p-value | $\beta$ [95% CI] | p-value |
| **<u>State dynamics</u>** |  |  |  |  |  |  |  |  |
| **Total Transitions** | 54.9 (18.7)<sup>MCI,CI</sup> | 48.0 (22.3) | 43.3 (21.6)<sup>HC</sup> | 42.1 (23.4)<sup>HC,CP</sup> | 4.92 | **0.002\*** | -5.78 [-11.23, -0.32] | **0.038\*** |
| **Fractional occupancy** |  |  |  |  |  |  |  |  |
| *State 1* | 0.11 (0.07)<sup>MCI,CI</sup> | 0.08 (0.06)<sup>HC</sup> | 0.07 (0.06)<sup>HC</sup> | 0.06 (0.06)<sup>HC,CP</sup> | 9.38 | **<0.001\*** | -0.02 [-0.04, -0.01] | **0.004\*** |
| *State 2* | 0.03 (0.03) | 0.02 (0.03) | 0.02 (0.03) | 0.02 (0.03) | 2.53 | 0.226 |  |  |
| *State 3* | 0.09 (0.05) | 0.08 (0.05) | 0.08 (0.05) | 0.08 (0.06) | 0.25 | 1.000 |  |  |
| *State 4* | 0.77 (0.10)<sup>a</sup> | 0.81 (0.10)<sup>HC</sup> | 0.83 (0.11)<sup>HC</sup> | 0.83 (0.10)<sup>HC</sup> | 5.55 | **0.004\*** | 0.03 [0.00, 0.05] | 0.053 |
| **Transition Probability** |  |  |  |  |  |  |  |  |
| *State 1 persist (NA=14)* | 0.29 (0.18)<sup>MCI,CI</sup> | 0.27 (0.18)<sup>CI</sup> | 0.22 (0.18)<sup>HC</sup> | 0.20 (0.20)<sup>HC,CP</sup> | 4.34 | **0.020\*** | -0.06 [-0.11, -0.02] | **0.007\*** |
| *State 1 → State 4 (NA=14)* | 0.57 (0.20) | 0.62 (0.21) | 0.63 (0.25) | 0.70 (0.24)<sup>HC,CP</sup> | 4.05 | **0.030\*** | 0.08 [0.02, 0.14] | **0.005\*** |
| *State 4 persist* | 0.84 (0.07)<sup>a</sup> | 0.86 (0.07)<sup>HC</sup> | 0.88 (0.07)<sup>HC</sup> | 0.88 (0.08)<sup>HC</sup> | 4.62 | **0.014\*** | 0.02 [0.00, 0.04] | 0.062 |
| *State 4 → State 1* | 0.08 (0.05)<sup>CI</sup> | 0.07 (0.05)<sup>CI</sup> | 0.05 (0.04) | 0.05 (0.04)<sup>HC,CP</sup> | 7.99 | **<0.001\*** | -0.02 [-0.03, -0.01] | **0.004\*** |
| | HC (N=59) | CP (N=131) | MCI (N=40) | CI (N=54) | F | p-value | $\beta$ [95% CI] | p-value |
| **<u>Control Energy</u>** |  |  |  |  |  |  |  |  |
| **Total Control Energy** | 0.09 (0.57)<sup>MCI,CI</sup> | 0.38 (0.73)<sup>MCI,CI</sup> | 0.66 (1.23)<sup>HC,CP</sup> | 0.57 (0.88)<sup>HC,CP</sup> | 6.56 | **<0.001\*** | 0.25 [0.04, 0.46] | **0.018\*** |
| *Persistence Energy* | 0.10 (0.73) | 0.35 (0.92) | 0.64 (1.03) | 0.53 (0.94) | 2.99 | 0.061 |  |  |
| *Transition Energy (NA=2)* | 0.08 (0.58)<sup>MCI,CI</sup> | 0.24 (0.76)<sup>MCI,CI</sup> | 0.62 (1.35)<sup>HC,CP</sup> | 0.58 (1.00)<sup>HC,CP</sup> | 6.19 | **<0.001\*** | 0.32 [0.09, 0.54] | **0.007\*** |
| **Transition Probability** |  |  |  |  |  |  |  |  |
| *State 1 persist (NA=93)* | 0.00 (1.00) | 0.05 (1.07) | 0.28 (1.84) | 0.01 (1.21) | 0.53 | 1.000 |  |  |
| *State 1 → State 4 (NA=17)* | 0.00 (1.00) | 0.12 (1.06) | 0.40 (1.53) | 0.47 (1.38) | 2.24 | 0.332 |  |  |
| *State 4 persist* | 0.00 (1.00) | 0.38 (1.07) | 0.66 (1.18) | 0.47 (1.03) | 3.30 | 0.084 |  |  |
| *State 4 → State 1 (NA=15)* | 0.00 (1.00)<sup>MCI,CI</sup> | 0.11 (0.93)<sup>MCI,CI</sup> | 0.64 (1.75)<sup>HC,CP</sup> | 0.51 (1.64)<sup>HC,CP</sup> | 3.87 | **0.038\*** | 0.34 [0.00, 0.67] | **0.049\*** |

*Note.* The total frequency of transitions was lower in cognitively impaired (CI) patients compared to preserved (CP) patients and healthy controls (HCs). CI patients spent relatively less time in *State 1* and more in *State 4*, with transition probabilities highlighting a general pattern of more transitions towards *State 4* and away from *State 1*. The total control energy and particularly the control energy associated with transitions was elevated in CI and mildly cognitively impaired (MCI) patients compared to CP patients and HCs. This was most notable for the transition from *State 4* to *State 1*. If a participant did not access a particular state this occasionally resulted in missing values (NA; i.e. not available). The reported p-values for the main group effects were corrected for multiple comparisons using Bonferroni and pairwise comparisons are reported if the corrected $p < 0.05$ (<sup>a</sup>=different from all other groups, <sup>HC</sup>=different from HC, <sup>CP</sup>=different from CP, <sup>MCI</sup>=different from MCI, <sup>CI</sup>=different from CI), \* $p < 0.05$ .

#### Fractional occupancy

After correcting for performing comparisons across four states, the fractional occupancy of *State 1* was significantly lower in CI compared to CP patients and HCs. Both MCI and CP patients also showed a lower fractional occupancy in *State 1* compared to HCs. For *State 2* and *State 3* the same directionality was observed, but no significant differences were found between groups. The fractional occupancy of *State 4* showed the opposite effect, being higher in CI patients compared to HCs. Although it was not significantly elevated in CI compared to CP patients, all MS groups showed a higher fractional occupancy in *State 4* compared to HCs. These findings indicate that the time spent in *State 1* and *State 4* were altered in MS, with CI patients mainly spending less time in *State 1*.

#### Transition probabilities

Based on the results above, the probability of persisting within and transitioning between *State 1* and *State 4* were explored further, correcting for performing four comparisons. Different probabilities of persisting in *State 1* were observed. CI persisted in *State 1* less frequently compared to CP patients and HCs. MCI also persisted in *State 1* less frequently compared to HCs. Differences between groups were also observed in the probability of persisting in *State 4*, but these were not observed between patients and only a heightened persistence probability was observed compared to HCs across all patient groups. Differences between groups were also observed in the probability of transitioning from *State 1* to *State 4*. When in *State 1*, CI transitioned to *State 4* more compared to CP patients and HCs. The inverse was true when in *State*, as then CI transitioned less to *State 1* compared to CP.

### Control energy

#### Total control energy

Of all 330 patients, 5 had incomplete diffusion MRI data (3 CP & 2 MCI) and were excluded from these analyses. Total control energy was increased in MCI and CI compared to CP patients and HCs (see Figure 4 & Table 2 for statistical parameters).

**Figure 4.**
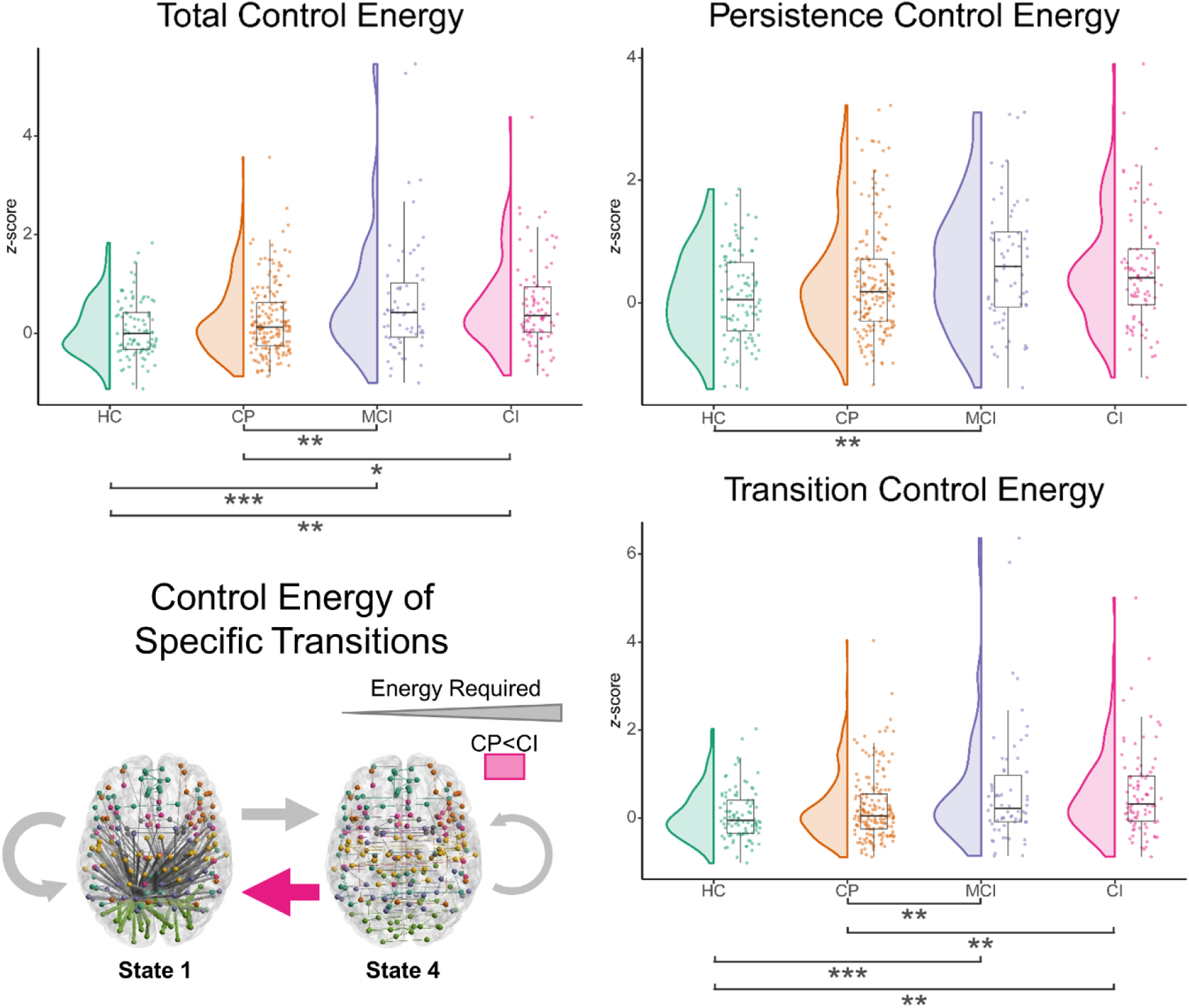
Control energy of brain state transitions across cognitive groups. In CI patients, transitions between and within brain states were more energetically costly compared to CP and HCs. In particular, more control energy was required for transition between states, not for persisting in the same state. The transition from *State 4* to *State 1* was particularly more costly in CI compared to CP patients. The thickness of the arrows on the bottom left indicates how much control energy is required on average in HCs for that transition. *Note. HC=healthy control, CP=cognitively preserved, MCI=mildly impaired, CI=cognitively impaired, *p<0.05, **p<0.01, ***p<0.001*.

#### Persistence and transition control energy

Persistence control energy was not different between groups after correction for performing two comparisons, thus staying in the same state was not more energetically costly in CI. Transition control energy was increased in MCI and CI compared to CP patients and HCs. This indicated that CI featured more energetically costly transitions between states. Based on these findings, we used transition and not total control energy in the correlations with disease severity.

#### Energy of specific state transitions

Similarly to the transition probability, the minimum control energy required for transitions between and within *State 1* and *State 4* were compared between groups, whilst correcting for comparing across these four transitions. Only the control energy of the transition from *State 4* to *State 1* showed significant differences between groups, with CI and MCI patients both showing increased control energy compared to CP patients and HCs. Control energy of transitions from *State 1* to *State 4*, as well as the control energy related to persisting in *State 1* or *State 4* did not differ between groups.

### Correlations with disease severity

#### Structural damage

We corrected significance thresholds for considering four measures of structural damage. Reduced white matter integrity in MS patients related to less frequent state transitions, an increased probability of persisting in *State 4*, more transition control energy, and more control energy for transitions from *State 1* to *State 4* (see Table 3 for statistical parameters).

**Table 3.** Correlational analyses.

|  | Total transitions<br>(N=330) |  | Transition Control Energy<br>(N=323) |  | *State 1 persist* |  |  |  | *State 1 → 4* |  |  |  | *State 4 persist* |  |  |  | *State 4 → 1* |  |  |  |
| --- | --- | --- | --- | --- | --- | --- | --- | --- | --- | --- | --- | --- | --- | --- | --- | --- | --- | --- | --- | --- |
|  |  |  |  |  | Probability<br>(N=316) |  | Control Energy<br>(N=243) |  | Probability<br>(N=316) |  | Control Energy<br>(N=308) |  | Probability<br>(N=330) |  | Control Energy<br>(N=325) |  | Probability<br>(N=330) |  | Control Energy<br>(N=310) |  |
|  | *r* | *p* | *r* | *p* | *r* | *p* | *r* | *p* | *r* | *p* | *r* | *p* | *r* | *p* | *r* | *p* | *r* | *p* | *r* | *p* |
| **Structural damage** |  |  |  |  |  |  |  |  |  |  |  |  |  |  |  |  |  |  |  |  |
| Cortical GM volume | 0.13 | 0.085 | -0.08 | 0.689 | 0.04 | 1.000 | -0.12 | 0.268 | -0.03 | 1.000 | -0.08 | 0.646 | -0.14 | 0.057 | -0.05 | 1.000 | 0.02 | 1.000 | -0.08 | 0.660 |
| Deep GM volume | 0.10 | 0.256 | -0.02 | 1.000 | 0.06 | 1.000 | -0.03 | 1.000 | -0.10 | 0.328 | -0.03 | 1.000 | -0.10 | 0.324 | 0.03 | 1.000 | 0.03 | 1.000 | -0.01 | 1.000 |
| Lesion volume | -0.07 | 0.925 | 0.00 | 1.000 | -0.03 | 1.000 | 0.03 | 1.000 | 0.01 | 1.000 | 0.03 | 1.000 | 0.07 | 0.967 | -0.04 | 1.000 | -0.06 | 0.993 | 0.04 | 1.000 |
| WM integrity | **0.16** | **0.013\*** | **-0.15** | **0.023\*** | 0.05 | 0.188 | 0.05 | 1.000 | -0.08 | 0.381 | **-0.16** | **0.005\*** | **-0.15** | **0.007\*** | -0.07 | 0.666 | 0.09 | 0.312 | -0.09 | 0.245 |
| **Clinical outcomes** |  |  |  |  |  |  |  |  |  |  |  |  |  |  |  |  |  |  |  |  |
| Average cognition | **0.15** | **0.023\*** | **-0.17** | **0.007\*** | **0.18** | **0.001\*** | -0.05 | 1.000 | **-0.17** | **0.002\*** | -0.12 | 0.062 | **-0.18** | **0.001\*** | -0.10 | 0.134 | **0.20** | **<0.001\*** | **-0.17** | **0.001\*** |
| EDSS | **-0.16** | **0.015\*** | 0.09 | 0.277 | -0.12 | 0.081 | -0.01 | 1.000 | **0.15** | **0.019\*** | **0.17** | **0.012\*** | **0.15** | **0.019\*** | 0.08 | 0.452 | -0.13 | 0.069 | 0.13 | 0.077 |
| Fatigue | -0.10 | 0.618 | 0.08 | 0.988 | -0.16 | 0.062 | 0.01 | 1.000 | **0.18** | **0.026\*** | 0.16 | 0.078 | 0.10 | 0.456 | 0.04 | 1.000 | -0.15 | 0.103 | 0.08 | 0.836 |
| **Cognitive domains** |  |  |  |  |  |  |  |  |  |  |  |  |  |  |  |  |  |  |  |  |
| Executive functioning | 0.08 | 1.000 | -0.12 | 0.302 | 0.09 | 0.457 | 0.06 | 1.000 | -0.11 | 0.256 | -0.12 | 0.120 | -0.10 | 0.280 | 0.00 | 1.000 | 0.12 | 0.105 | **-0.16** | **0.018\*** |
| Verbal memory | 0.07 | 1.000 | -0.03 | 1.000 | 0.08 | 0.933 | -0.12 | 0.265 | -0.11 | 0.194 | -0.01 | 1.000 | -0.09 | 0.524 | -0.07 | 1.000 | 0.10 | 0.284 | -0.01 | 1.000 |
| Processing speed | 0.10 | 0.568 | **-0.18** | **0.013\*** | **0.14** | **0.035\*** | -0.04 | 1.000 | -0.12 | 0.114 | -0.12 | 0.111 | -0.13 | 0.074 | -0.08 | 0.804 | 0.13 | 0.055 | -0.13 | 0.069 |
| Verbal fluency | 0.07 | 1.000 | **-0.18** | **0.015\*** | 0.12 | 0.169 | -0.12 | 0.271 | -0.09 | 0.628 | -0.10 | 0.440 | -0.10 | 0.318 | -0.02 | 1.000 | 0.12 | 0.097 | **-0.14** | **0.050\*** |
| Visuospatial memory | 0.03 | 1.000 | -0.05 | 1.000 | **0.17** | **0.007\*** | 0.02 | 1.000 | -0.10 | 0.286 | -0.03 | 1.000 | -0.07 | 1.000 | -0.10 | 0.374 | 0.12 | 0.150 | **-0.14** | **0.047\*** |
| Working memory | 0.02 | 1.000 | -0.08 | 1.000 | 0.11 | 0.259 | 0.11 | 0.349 | -0.05 | 1.000 | -0.02 | 1.000 | -0.07 | 1.000 | -0.04 | 1.000 | 0.12 | 0.143 | -0.09 | 0.604 |
| Attention | 0.08 | 1.000 | -0.05 | 1.000 | **0.16** | **0.013\*** | -0.10 | 0.500 | **-0.17** | **0.007\*** | -0.04 | 1.000 | -0.11 | 0.217 | -0.07 | 0.983 | 0.13 | 0.051 | -0.09 | 0.626 |

*Note.* These correlational analyses were performed on data from MS patients (N=330), with missing values for WM integrity (5), fatigue (163), executive functioning (12) verbal memory (2), IPS (2), verbal fluency (1), working memory (12), and attention (12). All correlations were adjusted for age, sex and level of education. The *p*-values were corrected for performing multiple comparisons using Bonferroni. GM=gray matter, WM=white matter, BPF=brain parenchymal fraction, EDSS=expanded disability status scale, \**p*<0.05.

#### Clinical outcomes

After correcting for investigating three clinical outcomes, worse average cognitive performance in MS related to fewer transitions and increased transition control energy. Further, average cognition related to all transition probabilities between *State 1* and *State 4*, whereas it only related to the control energy of transitions from *State 4* to *State 1*. Worse EDSS related to fewer transitions, a higher probability to transition from *State 1* to *State 4,* increased probability of persisting in *State 4*, and more control energy for transitions from *State 1* to *State 4*. More fatigue related to an increased probability to transition from *State 1* to *State 4*.

#### Cognitive domains

We corrected for studying seven cognitive domains. Worse information processing speed and verbal fluency associated with elevated transition control energy in MS. Impaired information processing speed, visuospatial memory, and attention related to less probability of persisting in *State 1*. Patients with poorer attention were more likely to transition from *State 1* to *State 4*. Finally, poorer performance on executive functioning, verbal fluency, and visuospatial memory related to more control energy required to transition from *State 1* to *State 4*.

## Discussion

This study showed that dynamic network changes required for normal cognitive processing requires more effort in cognitively impaired people with MS, as transitions between connectivity states required more control energy. This suggests that state transitions became more effortful and may explain why transitions happen less frequently in MS patients with cognitive impairment. The results showed that impaired patients spent more time in the *weakly connected state* and less in the *visual network state*, with the probability of transitioning to and from the *visual network state* being altered in CI patients. Transitions that happened more frequently did not become less energetically costly, but patients that required more transition control energy generally transitioned towards the *weakly connected state* and away from the *visual network state*.

CI patients used more control energy for transitions between connectivity states. This provides a mechanistic understanding of reduced brain dynamics in MS patients with cognitive impairment, suggesting that state transitions became more cognitively effortful and metabolically demanding,^20^ which may prevent transitions from occurring. Additional mediation analyses (see eMethods 2) did not support a clear directionality in which structural aberrations affect the transition probability; thus, it is possible that transitions occurring less frequently make them more effortful. Further experimental studies are required to discern this directionality. Several possible explanations exist for the more energetically costly network dynamics in CI MS. Firstly, demyelination-related conduction delays in MS can impact efficient information transfer,^38^ which matches our observed link with white matter integrity. We detected no relationship with lesion volume or atrophy in MS. This suggests that the extent of demyelination could affect these measures more strongly than changes in brain morphometry, which can be further studied using modelling work with individualized estimations of conduction velocities.^39^ Secondly, more energetically costly state transitions may be linked to an excitation-inhibition imbalance,^40^ as an adequate balance is critical for efficient neural encoding.^41^ This matches a recent study showing the relevance of such balance for cognition in MS.^42^ Thirdly, structural damage in MS might impair the efficient wiring of the structural network and make functional transitions more energetically costly.^43^ Exploratory analyses using a “healthy” structural network to compute control energy showed that differences between groups remained (see eMethods 3). Thus, although the structural network likely shaped network dynamics, this suggests that he dynamic patterns themselves might particularly be more costly and not the organization of the structural network.

The *visual network state* was especially important for cognition, as CI-MS was less likely to stay in this state and more likely to move to the *weakly connected state* than CP patients. Conversely, when in the *weakly connected state*, CI patients moved to the *visual network state* less. Thus, in contrast to our hypothesis, CI patients did not get “stuck” in states featuring high DMN connectivity (the *weakly connected state*), but also returned to that state more often. These transition probabilities could not be explained by the control energy required for the transitions, as exploratory analyses suggested that a less energetically efficient network pushes the brain to the *weakly connected state* (see eMethods 4). This *weakly connected state* was by far the most abundant across participants, which was further elevated in CI patients. Lower connectivity strength is arguably less metabolically demanding,^16^ so residing in this state more regularly could be a compensatory mechanism. These periods of low connectivity may also be uniquely relevant for cognition,^18^ but were likely underrepresented in studies utilizing static or windowed approaches,^16^ emphasizing the utility of framewise approaches. Periods of more integrative connectivity (the *visual network state*) were observed less in CI patients, whereas instances of more integrated processing are likely important for complex cognitive tasks^44^ which is often impaired in MS.^1^ Finally, hampered transitions from a state featuring more DMN connectivity (the *weakly connected state*) to the *visual network state* might reflect disrupted switching between internally and externally oriented processing, as previously proposed.^12^

We found a link between transition frequency and probability with total cognition. The frequency of transitions across all states may be a broad indicator of disease severity, however, as we also observed that fewer transitions related to more physical disability. This aligns with findings from prior research on shifts in connectivity states in MS.^15,45,46^ The same might be true for the detected increase in control energy for CI patients, as recent work showed that MS patients with physical disability required more control energy compared to non-disabled patients.^21^ In contrast to that finding, we did not observe a relationship between physical disability and transition control energy in this study, possibly as our cohort had longer disease durations where disability mechanisms might be different. Interestingly, the transition control energy was particularly relevant for information processing speed and verbal fluency which suggests that transition control energy might be related to shared cognitive processes, such as cognitive flexibility. Altered network dynamics could in theory impact fatigue in MS, given the previously observed link between fatigue and energy metabolism.^47^ We did not observe such a relationship in the current study, so dedicated studies now need to test whether altered dynamics are important for specific types of fatigue.

The current framework comes with inherent strengths and challenges. Network control theory provides an exciting new perspective to link brain function to structural network organization. Although control energy should not be directly equated to metabolic energy, previous work did show a relationship between control energy and glucose metabolism.^43^ Parameter choice is still a topic of debate, however, particularly for the arbitrary choice of a control horizon, which is why we optimized this parameter using a data-driven approach (see eMethods 1).^21^ Despite these challenges, the current framework offers an exciting opportunity and generalizable approach to study and develop personalized treatment of cognitive impairment in MS by identifying optimal target areas for transcranial magnetic stimulation or for tailoring pharmacological interventions.^48,49^ Furthermore, although fine-grained temporal scales can increase noisiness of windowed connectivity (i.e., short windows), the current approach utilizes information (e.g. variance) from the entire scan and is not affected by noisy estimations in the same way.^17^ Furthermore, the chance of sojourning in the same connectivity state was particularly low for event states (i.e., brief high-amplitude co-fluctuations), so to understand what is happening during those states other modalities that acquire data with a better temporal resolution (e.g. electro-/magnetoencephalography) are required. Finally, explicit stimuli (e.g. tasks or movie watching) would be required to understand the cognitive processes underlying specific states.^50^

## Conclusions

This study showed that transitions between connectivity states became more effortful in MS patients with cognitive impairment compared to preserved patients and controls, as these were more costly from a control energy perspective. Heightened energetic costs might limit the transitions between states and, in turn, negatively impact cognition. The transitions between a *weakly connected state* and a slightly more integrated *visual network state* appear to be particularly relevant for cognition in these patients. These findings provide new insights into the possible biological underpinnings of disturbed brain dynamics in CI patients with MS and opens up new avenues for treatment of cognitive impairment in MS.

## Data availability

Anonymized data, not published in the article, will be shared on reasonable request from a qualified investigator. Code is available on GitHub: https://github.com/taabroeders/research-projects/tree/main/states_2024.

## CRediT authorship contribution statement

**T.A.A. Broeders:** Conceptualization, Formal analysis, Methodology, Visualization, Writing - original draft, Writing – review & editing. **M. van Dam:** Methodology, Writing – review & editing. **G. Pontillo:** Methodology, Writing – review & editing. **V. Rauh:** Conceptualization, Writing – original draft. **L. Douw:** Writing – review & editing. **Y.D. van der Werf:** Supervision, Writing – review & editing. **J. Killestein**: Writing – review & editing. **F. Barkhof**; Writing – review & editing. **C.H. Vinkers:** Supervision, Writing – review & editing. **M.M. Schoonheim:** Conceptualization, Methodology, Resources, Supervision, Writing – review & editing.

## Supplementary material

Supplementary material is available at *Neurology* online.

## Supporting information

eMethods

