## Supplementary material for "Energetically costly functional network dynamics in cognitively impaired multiple sclerosis patients": eMethods

**eMethods 1. Network control theory and minimum control energy**

**eMethods 2. Mediation analysis**

**eMethods 3. Control energy with a healthy structural network**

**eMethods 4. Correlating transition probability with control energy**

**eReferences**

### eMethods 1. Network control theory and minimum control energy

To use control theory and determine the control energy required to transition between connectivity states, the ′nctpy′ Python toolbox was used. The change of brain activity (fMRI signal) over time ($\dot{x}$) was determined by a 'natural trajectory' and some 'external input'. The natural trajectory described the evolution of activity relative to the activity observed at time *t* ($x_t$) constrained by the structural pathways (normalized number of streamlines) and a global signal decay ($A$). The external input is "extra" activity, or *control energy*, that was infused in the system ($u$), which was not confined to any particular set of brain regions ($B$; i.e., $B$ was an identity matrix). Thus, the temporal evolution of activity was modelled by: $\dot{x} = Ax(t) + Bu(t)$.

To determine the *minimum control energy*, the controllability Gramian ($W$): $W = \int_0^T e^{At} BB^\top e^{A^\top t} dt$ was computed. The control horizon ($T$) characterizes the number of steps that activity can disperse across anatomical connections. The optimal value for $T$ was determined by computing the minimum control energy for $T$=[0.501-2.501] in steps of 0.5 and searching for the strongest anti-correlation with the observed probability of transitions, as suggested previously.[1] This resulted in an optimal $T$ of 1.501. The minimum control energy was calculated by: $E_{min}^R = (e^{AT}x0 - xt)^\top W^{-1}(e^{AT}x0 - xt)$. The result was one $E_{min}^R$ value for each brain region and frame, whereas the sum over all regions determined overall energy required for that transition ($E_{min}$).

#### eMethods 2. Mediation analysis

We expected that a relationship between control energy and cognition would be mediated by the frequency of transitions, which was based on the intuition that more energetically costly and, thus, effortful transitions would make transitions happen less. The resultant restriction in functional brain network dynamics might, in turn, impair cognition. Using data from MS patients, the pairwise relationships between total transition frequency, transition control energy and cognitive impairment were determined. If all three were significant, the change in predictive value of control energy for cognition was determined when the transition frequency was added to the model, with $\beta$-change>10% indicating a significantly mediating effect.[2]

MS patients with a higher transition control energy generally showed a lower number of transitions (A→B; $\beta$=-10.114±1.114, $p$<0.001). A lower number of transitions in these patients related to lower average cognition (B→C; $\beta$=0.006±0.002, $p$=0.008). More transition control energy related to more impaired cognition (A→C; $\beta$=-0.158±0.052, $p$=0.002). The strength of this last relationship reduced when controlling for the number of transitions (A→B→C; $\beta$=-0.114±0.058, $p$=0.048, $\beta$-change=27.8%), suggesting that the link between transition control energy and average cognition was mediated by the number of state transitions. We also observed this inverse mediating relationship (B→A→C; $\beta$=-0.004±0.003, $p$=0.095, $\beta$-change=29.4%), so care is warranted when interpreting directionality.

#### eMethods 3. Control energy with a healthy structural network

We aimed to additionally disentangle whether inefficient connectivity state transitions are directly related to structural network disorganization or can be observed by the functional pathways (i.e., brain network dynamics) alone. Therefore, we computed transition control energies again, but this time with an "healthy" structural network. To that end, the structural connectivity matrices were averaged across controls and the resulting mean connectivity matrix was normalized. This normalized structural matrix was used for the computation of minimum control energy across all participants. Then, the same procedure was used to compute transition control energy as for the main analysis, including the same statistical comparisons.

Similarly to the original finding, the resulting transition control energy ($F(3,411)=6.413$, $p<0.001$), was notably increased in MCI and cognitively impaired CI compared to CP patients (MCI: $\beta=0.371$, 95%-CI=[0.12, 0.62], $p=0.004$; CI: $\beta=0.319$, 95%-CI=[0.09, 0.55], $p=0.007$) and controls (MCI: $\beta=0.499$, 95%-CI=[0.22, 0.78], $p<0.001$; CI: $\beta=0.446$, 95%-CI=[0.19, 0.71], $p<0.001$).

#### eMethods 4.Correlating transition probability with control energy

We wanted to additionally explore the linear relationship between state transition probabilities and the associated control energy requirements in MS patients. To do this, we computed Pearson's correlations between for the total transition frequency or transitions probabilities for *State 1* and *State 4* on the one hand, with transition control energy and the associated transition-specific control energy measures on the other hand.

We corrected for performing five correlations, including the total transitions and four specific transitions. In MS, more transition control energy related to a lower number of total state transitions ($r(321)=-0.487$, $p<0.001$), meaning that if transitions are more energetically costly they tend to happen less frequently. With regard to specific transitions, control energy associated with persisting in *State 1* did not correlate with the relative probability of persisting in that state ($r(241)=-0.080$, $p=0.861$). More control energy associated with persisting in *State 4* correlated to a heightened probability of persisting in that state ($r(323)=0.680$, $p<0.001$). Similarly, more control energy required for transitioning from *State 1* to *State 4* related to an increased relative probability of that transition occurring ($r(306)=0.414$, $p<0.001$). The inverse transition, from *State 4* to *State 1*, showed an inverse relationship, with more control energy being related to fewer transitions ($r(308)=-0.304$, $p<0.001$).
